# A municipality-based approach using commuting census data to characterise the vulnerability to influenza-like epidemic: the COVID-19 application in Italy

**DOI:** 10.1101/2020.05.12.20100040

**Authors:** L. Savini, L. Candeloro, P. Calistri, A. Conte

**Affiliations:** National Reference Centre for Veterinary Epidemiology, Programming, Information and Risk Analysis. Istituto Zooprofilattico Sperimentale dell’Abruzzo e Molise “G. Caporale”, Teramo, Italy

**Keywords:** COVID-19, commuting census data, municipality-specific contact rate, vulnerability, infectious disease modelling

## Abstract

In February 2020, Italy became the epicentre for COVID-19 in Europe and at the beginning of March, in response to the growing epidemic, the Italian Government put in place emergency measures to restrict the movement of the population. Human mobility represents a crucial element to be considered in modelling human infectious diseases.

In this paper, we examined the mechanisms underlying COVID-19 propagation using a Susceptible-Infected stochastic model (SI) driven mainly by commuting network in Italy. We modelled a municipality-specific contact rate to capture the disease permeability of each municipality, considering the population at different times of the day and describing the characteristic of the municipalities as attractors of commuters or places that make their workforce available elsewhere.

The purpose of our analysis is to provide a better understanding of the epidemiological context of COVID-19 in Italy and to characterize the territory in terms of vulnerability at local or national level. The use of data at such a high spatial resolution allows highlighting particular situations on which the health authorities can promptly intervene to control the disease spread.

Our approach provides decision-makers with useful geographically detailed metrics to evaluate those areas at major risk for infection spreading and for which restrictions of human mobility would give the greatest benefits, not only in the beginning of the epidemic but also in the last phase, when the risks deriving from the gradual lockdown exit strategies must be carefully evaluated.

## Introduction

On 31 December 2019, the Chinese Country Office of the World Health Organization (WHO) was informed about cases of pneumonia of unknown etiology in residents of Wuhan City, Hubei Province of China [1]. Later defined as a new disease (COVID-19) caused by a novel coronavirus (SARS-CoV-2), the epidemic was declared by WHO a public health emergency of international concern on 30 January and a “pandemic” on 11 March. In February 2020, Italy became the epicentre for COVID-19 in Europe and on the 22 February the Italian Government imposed a lockdown in hotspot areas in Lombardy and Veneto region [2,3]. On 9 March 2020, in response to the growing epidemic, emergency measures restricting the movement of the population (except for essential work categories and health reasons), were extended to the entire country.

Human mobility represents a crucial element to be considered in modelling human infectious diseases and the main factors influencing mobility patterns and magnitude depend on the considered scale (global, national, local). At global level, the study of air traffic connections may provide good indications for predicting the worldwide spread of a human infectious disease [4,5]. At a local scale other types of movement must be taken into account. Recently, various studies have been carried out to evaluate how different human mobility data, such as mobility data by Google or data collected through mobile phone, can guide government and public health authorities to evaluate the effectiveness of measures to control the COVID-19 spread [6,7]. However, commuting, defined as the daily movements from residence to work or school, is certainly the most relevant and widely studied factor to describe spatial mobility in local models [8–10].

In this paper, we analyse the commuting flows in Italy, using census data (ISTAT 2011) [11], in order to assess its contribution in spreading the COVID-19 at the beginning of the epidemic.

We first calculated Social Network Analysis (SNA) centrality measures at province level, and then we examined the underlying mechanisms of propagation using a stochastic Susceptible-Infected (SI) model mainly driven by commuting network and applied to a revised conceptualization of the contact rate parameter. We modeled a municipality-specific contact rate to capture the vulnerability to the disease of each municipality, considering the population in different times of the day and considering the characteristic of municipalities as attractor of commuters or displacing their workforce elsewhere.

The aim of our analysis is to provide a better understanding of the epidemiological context of COVID-19 in Italy, and to characterize the territory in terms of vulnerability either at local or national level. The objective is not to precisely estimate the exact number of cases or the magnitude of the epidemic in absolute manner, but to understand how the disease can spatially and temporally spread countrywide. In particular, we illustrated three scenarios: in the first scenario, the model estimates the disease transmission by calculating the expected number of cases and municipalities affected by the virus between 26 February and 6 March. We focused on the first ten days of the epidemic because most of the impact that commuting may have had is before the application of the lockdown measures. In the second scenario we explore and compare the spread patterns in three different regions (Lombardy, Abruzzi and Basilicata), which were differently affected by the COVID-19 spread. In scenario 3, the local spread in the Abruzzi region is simulated to define the vulnerability of each municipality to a resurgence of the COVID-19 epidemic during future partial or total easing of the lockdown measures.

This approach could help Health Authorities and policy makers to implement and direct the right interventions to contain the rapid expansion of the emergency. This contribution, although based on the current epidemic, will provide useful elements for other influenza-like epidemics that might happen in the future.

## Material and methods

### Demography and commuting Network

To analyse the Italian commuting network we used census data collected in 2011 [11]. All data is obtained at municipality level. About half of the 60 million people living in Italy declared a daily movement to their usual place of study or work.

After adjusting the geographical dataset of the Italian municipalities according to the modifications occurred after 2011, the matched commuting dataset contains 7,915 municipalities, a resident population of 60,340,328 and 28,805,440 commuters – within (17,497,742) and between (11,307,698) municipalities.

Commuting flows directed to or coming from abroad are not considered in the analysis. Figure 1 shows the distributions of the Italian population (a) and commuters (b).

**Figure 1.**
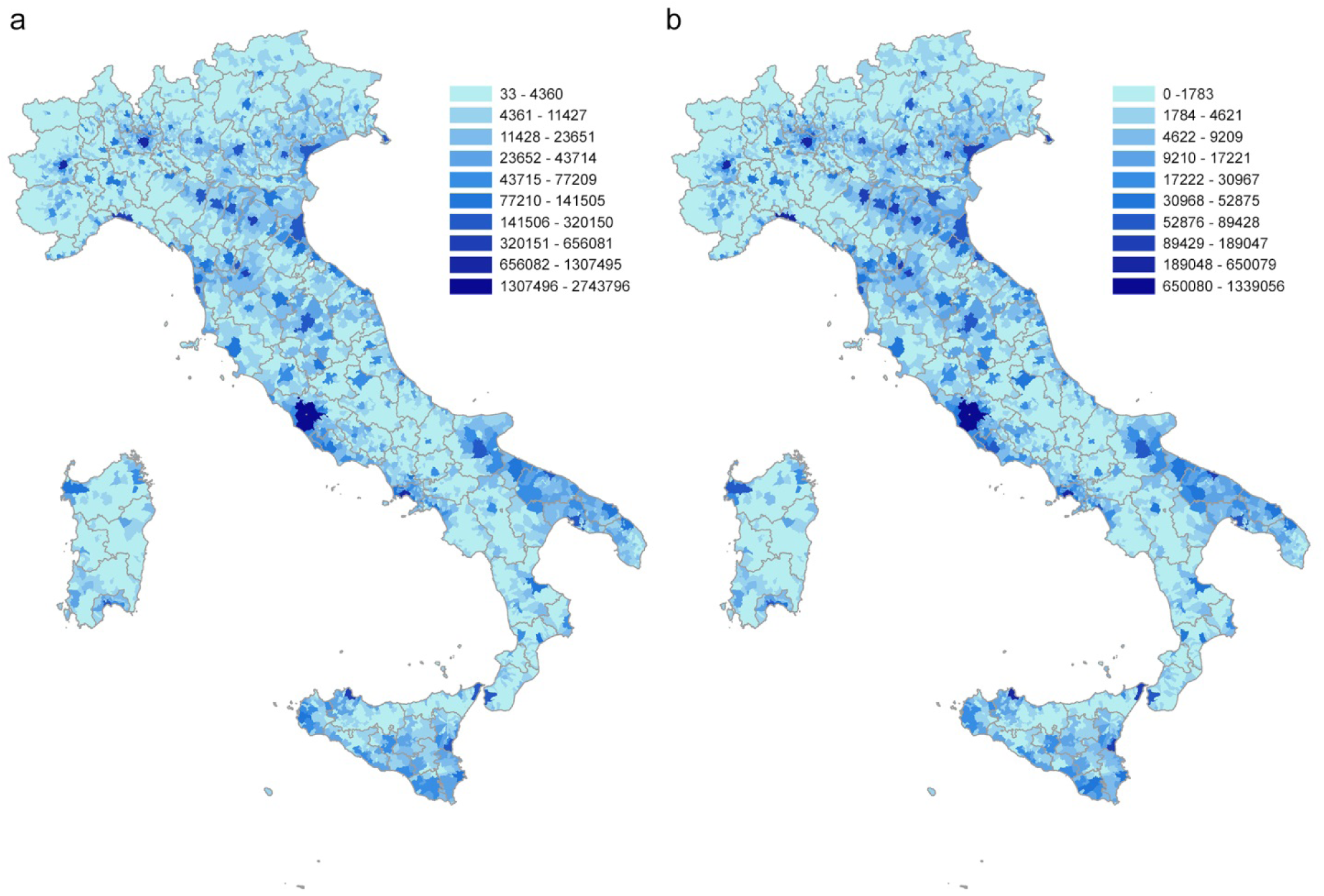
Distribution of the Italian population (a) and of daily commuters (b) at municipality level. Source ISTAT.

A commuting network is generated by creating a direct weighted edge between two nodes, represented by the municipalities of origin and destination. The weight indicates the number of commuters traveling on that connection in a typical working day (Figure 2a). The resulting network is composed by 7,915 nodes and 539,223 edges.

**Figure 2.**
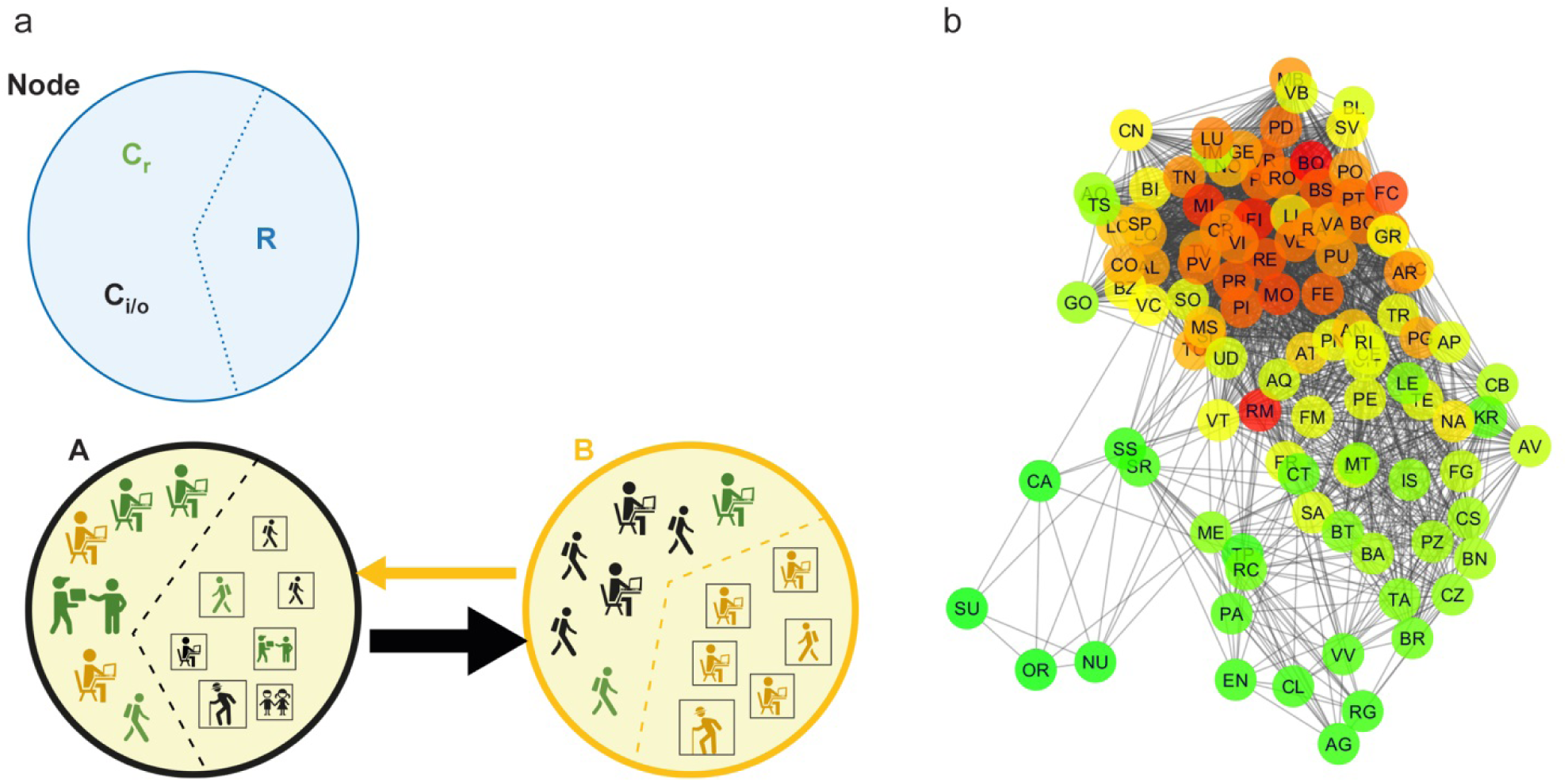
Commuting network structure. (a) All categories of commuters and non-commuters that characterize each node of the network on a typical working day are represented: commuters inside the node (**C_r_**), incoming and outgoing commuters (**C_i_** and **C_o_** respectively) and residents who are not commuters (**R**). The edge between two nodes, from a source to a destination, is represented by an arrow (direction) and its size is proportional to the number of commuters moving daily. (b) Graph representation of the commuting network at province level (undirected for displaying purpose). The scale colour from green to red is based on the node degree value.

Then, the commuting network is rescaled at province level (the lowest NUTS level - Nomenclature of Territorial Units for Statistics, NUTS 3) in order to compare the results of the network analysis with the COVID-19 cases as recorded at province level by the Dipartimento della Protezione Civile and archived on GitHub [12]. The rescaled network has a size of 107 nodes and 3,310 edges (Figure 2b).

The rescaled commuting network is analysed by calculating the centrality measures commonly used in epidemic modelling [13–15]: degree (in-out), strength (in-out), betweenness - both for the global network and for sub-networks (incoming and outgoing commuters greater than 50, 100 and 1000).

### SI model to evaluate the COVID-19 spread dynamics

#### Theoretical geo-demography framework

The model is based on commuting network at municipality level. In a municipality we have the resident population divided in: in non-commuting (R), commuting within the municipality (C_r_) and commuting outgoing the municipality (C_o_). Commuting affects the number of people present in a municipality during the different times of the day, significantly modifying the registered resident population. If we define C_i_ as the non-resident population commuting into a municipality, we have that the real population in a specific time of the day is given by R+C_r_-C_o_+C_i_ (Figure 2a).

In the right side of Figure 3 three municipalities (A, B, C) with their registered residents are shown; the left side shows the actual population present in the municipalities, following in and out commuting during the day. In municipality C, the resident population equal to 10, becomes 4 during the day, due to a prevalent component of C_o_. Conversely, the population in B significantly increases during the day compared to the resident population, due to the higher C_i_ component (from 10 to 13). Municipality A has a balanced population during the entire day, having an equivalent C_o_ and C_i_ components.

**Figure 3.**
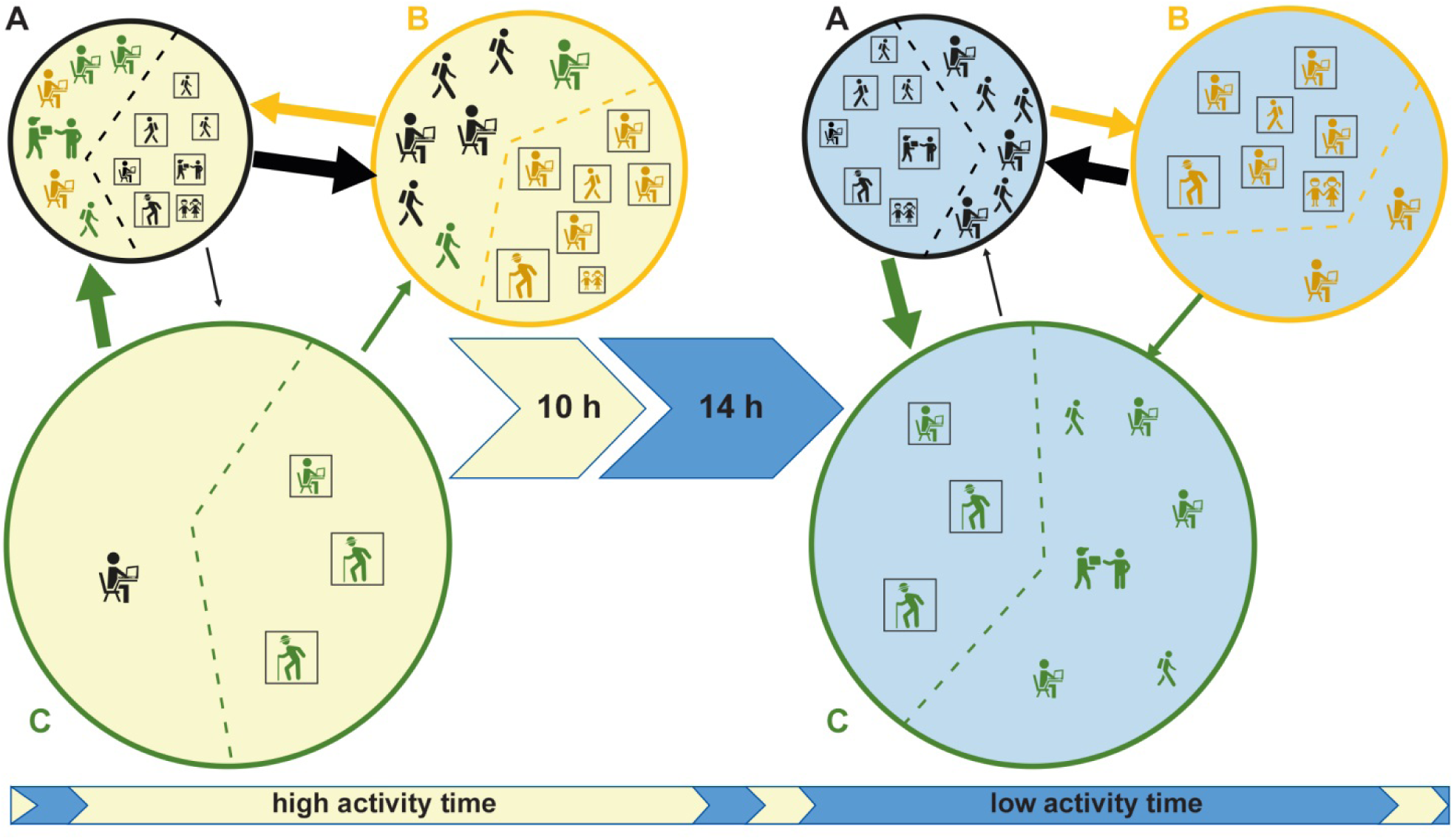
A typical working day description considering three different municipalities and populations. Population changes due to commuting in two different times of the day (high activity and low activity). The municipalities, A, B, C are displayed by circles in black, yellow, and green; the icons, coloured according to the circles, represent the resident populations in the 3 municipalities; squared icons represent non-commuters resident (R) and C_r_ component; the arrows represent commuters moving daily from a source to a destination (C_o_ and C_i_) and the arrow’s size is proportional to the number of moving commuters. The municipality C, with a population of 10, decreases to 4 during the day, due to a prevalent component of outgoing commuters (C_o_=7). Conversely, the population in B increases significantly during the day, due to the higher incoming commuters (C_i_ =7). Municipality A has a balanced population during the entire day, having an equivalent C_o_ and C_i_ components.

The typical working day is divided in two parts based on people’s daily contact dynamics: ‘high activity’ time during which contacts are facilitated by the social common activities (e.g. work, school, sports and similar) and ‘low activity’ time in which the activities are reduced or stopped (e.g. during night or sleep time).

The ‘high activity’ and ‘low activity’ times of the day, in combination with the resident and commuters populations, determine different levels of contact (and therefore of infection) between municipalities and between individuals within the municipality.

#### Model implementation

A Susceptible-Infectious (SI) compartmental model is implemented to simulate disease spread due to the commuting between municipalities, taking into account not only the absolute commuters values, but including the influence of the typical structure of a day (high and low activity) as described in the previous paragraph. It is an agent-based model where a subject susceptible can became infectious if living or working within an infected population. The following ODEs system describes the model:

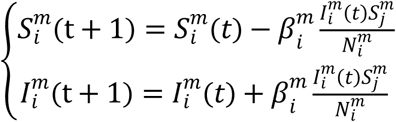

where S = susceptible population, I = Infected population, N population size, *β* is the infection contact rate, *i* indicates the municipality, *m* indicates the time of the day (high active or low active time) and *t* is a specific day; the equation describing the transition state of each individual, from susceptible (s) to infected (i), follows a Bernoulli distribution:

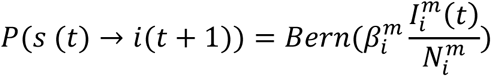

Therefore, we assume a homogeneous mixing of the population adopting two infection contact rates to take into account the time of the day (high active or low active time). The effective contact rate (β_0_ = 0.244) is estimated from the temporal evolution of the cases observed between 26 February and 6 March, assuming an exponential infection growth [16] and considering a basic reproductive number (R_0_) ranging from 2.24 to 5.71 [17].

#### Contact rate parameter modelling

We illustrate the approach used to calculate different β values for each municipality in different times of the day (low and high activity times).

The effective contact rate (β_0_ = 0.244) is estimated from the temporal evolution of the cases observed between 26 February and 6 March, assuming an exponential infection growth.

We use the following equation to scale the daily value of the infection contact rate:

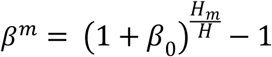

where *H* is the number of hours in which the hourly β is greater than zero (we assume the hourly β = 0 during sleeping time – 10 hours per day) and H_m_ is the number of hours relative to ‘high’ and ‘low’ activity times (H = 14 and H_m_ = 11 (during ‘high activity’ time), 3 (during ‘low activity’ time)). Thus, being β_0_=0.244, β^m^= {0.1871; 0.0479}.

However, in our model we define the contact rate for each municipality as depending on the population present in different times of the day:

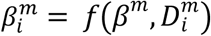

where *D* is the population density according to commuting flows in municipality *i* and time m.

However, assuming the same β for the whole country implies the disease spreads with the same strength everywhere, even inside those areas highly different spatially, demographically and in terms of commuting systems. So our approach takes into account different factors modulating the β value:

- β value differs during the day: in Figure 5, municipality C has a higher β during ‘low activity’ time in comparison to the ‘high activity’ one, whilst for B is the opposite. Municipalities like B can be considered as “attractors of commuters” having a high rate of incoming commuters. Municipalities like C, having a high rate of outgoing commuters are usually emptied during the working time.
- β value depends on the density of people staying in a municipality in a specific time of the day. However, we correct the population density so to be increased when the number of commuters is higher than the number of non-commuters and vice-versa. In Figure 3, municipality C has a daily β lower than B and A due to its extension (that reduces the density population). But its population density is higher during the ‘low activity’ time (due to commuters coming back) in comparison to the ‘high activity’ time. Therefore, the population density of C is raising due to the number of commuters coming back during the “low activity” time, whilst its population density remains unchanged during the ‘high activity’ time since its commuting components are equal to its non-commuters population.

The calculation of β for each municipality and time of the day is described in detail in Supplementary Information.

### Model Simulation Scenarios

In order to highlight the different perspectives of the developed model and the potential of its application, three scenarios are considered:

***Scenario 1***. COVID-19 spreading at municipality level for the entire Italian territory between 26 February and 6 March. This scenario is used to validate our model in relation to the current epidemic during the first 10 days of spread. The model assumes that no restrictions of human mobility are put in place at the beginning of the epidemic. Inside each province with confirmed COVID-19 cases at the starting time (26 February), the infected people are assumed to be randomly distributed. Results are then compared at province level with the official data on confirmed cases.
***Scenario 2***. Local spreading (during the first 21 days of the epidemic). This scenario is implemented to explore and compare the spread patterns in different regions (rather than validate the predictive capacity), to better understand how the human mobility as spread driver can affect the epidemic. Lombardy, Abruzzi and Basilicata regions are chosen based on the level of infection (high, medium and low) observed during the epidemic. In this scenario we assume that the infection starts at municipality level (the first infected municipality notified in the region is considered as seed); no restrictions of human mobility are put in place at the beginning of the epidemic; the regional network is closed to external commuting exchanges. A longer timeframe is chosen to better explore and compare the different spread patterns.
***Scenario 3***. Local spread in the Abruzzi region is simulated (during the first 14 days of the epidemic) considering each municipality as a seed for each simulation. This scenario aims at defining the vulnerability of each municipality to a new epidemic. The choice of the Abruzzi region has only an illustrative purpose. We assume that the infection starts in turn in each municipality so to assess the weak points of the whole region.

All simulations use the infection contact rate β described in the previous paragraph, assuming the number of plausible active cases (K) is ten times the number of officially detected active cases, as reported by the Italian Institute for International Political Studies [18]. In Table 1 all the simulation parameters used in the three scenarios are listed.

**Table 1.**
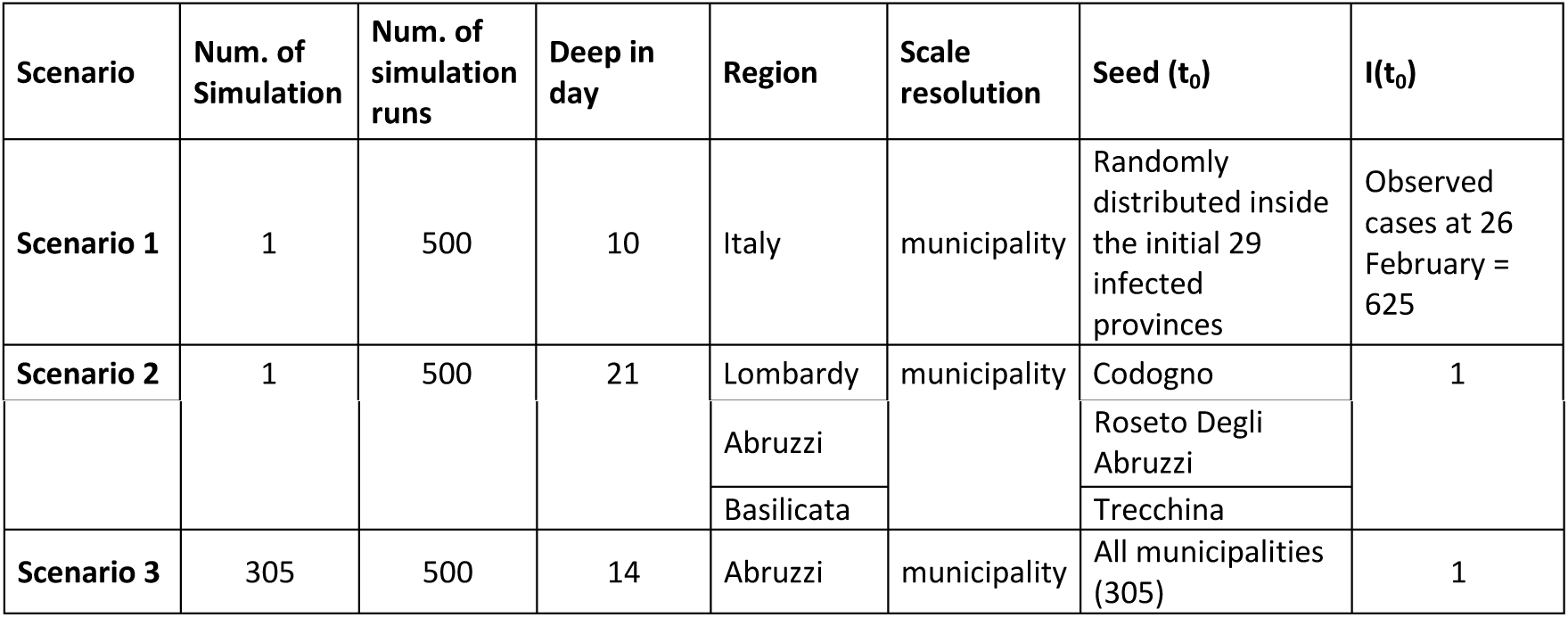
Simulation parameters for each scenario.

During the first five days of the week, as working days, the β is calculated considering the “high activity” time for the commuters adjusted population (R+C_r_-C_o_+C_i_), whereas the “low activity” time is applied to the resident population (R+C_r_+C_o_). The opposite occurs during the weekend (the last two days), to take into account that the resident population becomes more active than that determined by commuter flows.

The model is run at municipality level. For each node (municipality), for each individual of the node, and for each temporal step t of a day, the model recalculates the state (in terms of S and I) of the source and destination nodes for the next temporal step by following the equations above described.

The pseudo model algorithm is coded as follows:

*#constants shared between scenarios K=10*

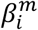

*#constants within scenario*

*SIM (number of simulation)*

*Deep (number of days to be simulated)*

*NTW (Commuting network: weighted links among municipalities of the corresponding scenario)*

*I (number of officially infected)*

*For each simulation*

*Sample infected according to scenario*

*For each day*

*If during weekend, switch the correspondence between active period and population For each sub-day period*

*For each infected municipality*

*find new infected according to ODE equations*

*End*

The model is stochastically implemented in R-software (Version 3.6, R-Foundation for Statistical Computing, Vienna, Austria); “doparallel” R package is used for parallelizing the simulations [19]. Figures are created in R using “sp” and “ggplot2” libraries [20,21], in ArcMap 10.5 ESRI and Cytoscape (Version 3.2.1) programs.

## Results

### Analysis of the commuting network

A Pearson’s correlation matrix was calculated among the SNA metrics of the commuting network (and sub-networks) and COVID-19 cases (at different times of the epidemic), (Figure 3a).

Degree measures and COVID-19 cases showed a significant correlation (p< 0.05). In particular, the degree calculated for the sub-network built on the basis of incoming and outgoing commuters greater than 50 (Deg50) and COVID-19 cases (on the 26 March 2020) have the highest correlation value equal to 0.72 (black bordered square in Figure 4a). Figure 4b represents a scatter plot between Deg50 and COVID-19 cases in logarithmic scale. To capture the commuting behaviour of each province (node) the scale colour from green to red is used to characterize the in-strength (incoming commuters from lower to higher) and the symbol size characterizes the node in terms of out-strength (outgoing commuters from smaller to bigger). It is evident that the northern provinces, most affected by the disease, are also those characterized by a high degree (flow of commuters among multiple provinces) and high strength (incoming and outgoing commuters exchanged) while the provinces of central and southern Italy, with a lower number of cases, are mainly characterized by lower degree and in-strength values.

**Figure 4.**
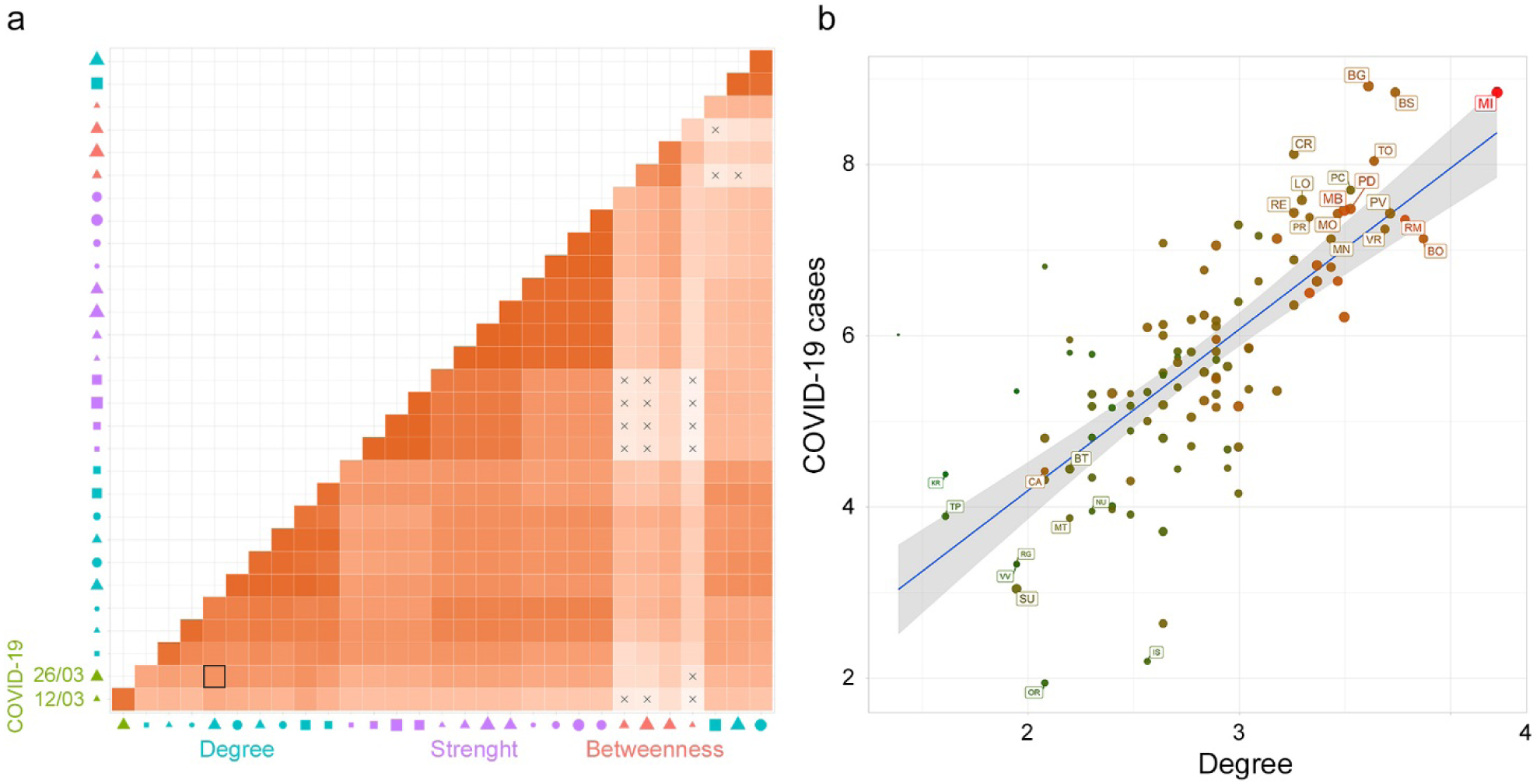
Correlation analysis. (a) The heatmap of Pearson’s correlation coefficients among all the variables: SNA metrics for the rescaled commuting network and sub-networks (with in and out commuters greater than 50, 100 and 1000), and COVID-19 cases (at different times). The X symbol indicates a non-significant correlation. The red scale colour indicates a positive Pearson’s coefficient value. In the x-axis legend, the symbol colour represents the variable groups: COVID-19 cases (green), betweenness (red), degree (light blue), and strength (purple); the symbols: circle, square and triangle represent the directions in terms of in (circle), out (square), and none (triangle); the symbol size from smaller to bigger represents the networks variables (sub-networks -with in and out commuters greater than 50, 100 and 1000 and the whole network).(b) A scatter plot graph between Deg50 and COVID-19 cases (on the 26^th^ of March 2020) in logarithmic scale. The scale colour from green to red is used to characterize the in-strength (from lower to higher number of incoming commuters) of the node and the symbol size (from smaller to bigger) characterizes the node in terms of out-strength (lower to higher number of outgoing commuters).

### Contact rate parameter estimation

The contact rate β values are calculated on the basis of commuting data and population size in each municipality. Figure 5a shows the β values per municipality (quantile aggregation), while Figure 5b shows the statistically significant hot spots, cold spots, and spatial outliers of β values using the Anselin Local Moran’s I statistic [22]. Although the map of contact rates mainly reflects the resident population density, the municipality-specific contact rate captures the disease permeability of each municipality, considering the population in different moment of the day and depicting the characteristic of municipalities as attractor of commuters or as diplacer of its workforce elsewhere. The clusterization of geographical areas with similar characteristics in terms of vulnerability to the introduction and spread of the disease is highlighted by Moran’s analysis (Figure 5b). It is evident that there are large areas characterized by the homogeneous presence of high β values (pink areas). In these areas, the introduction of the disease inevitably leads to a spread more difficult to control due to the simultaneous presence of connections between municipalities and the high population density. On the contrary, the large areas with low β levels (light blue) represent areas in which the disease spreads slowly (e.g. Alpine and Apennine areas, Basilicata, Sardinia, the southern part of Tuscany and Molise). The red areas constitute potential outliers which, despite a high β value, would hardly expand rapidly the disease in the surrounding areas which have instead low β values. In this way Figure 5b, grouping similar municipalities’ values, gives an immediate and overall picture of the Italian territory in terms of higher or lower susceptibility to an epidemic.

**Figure 5.**
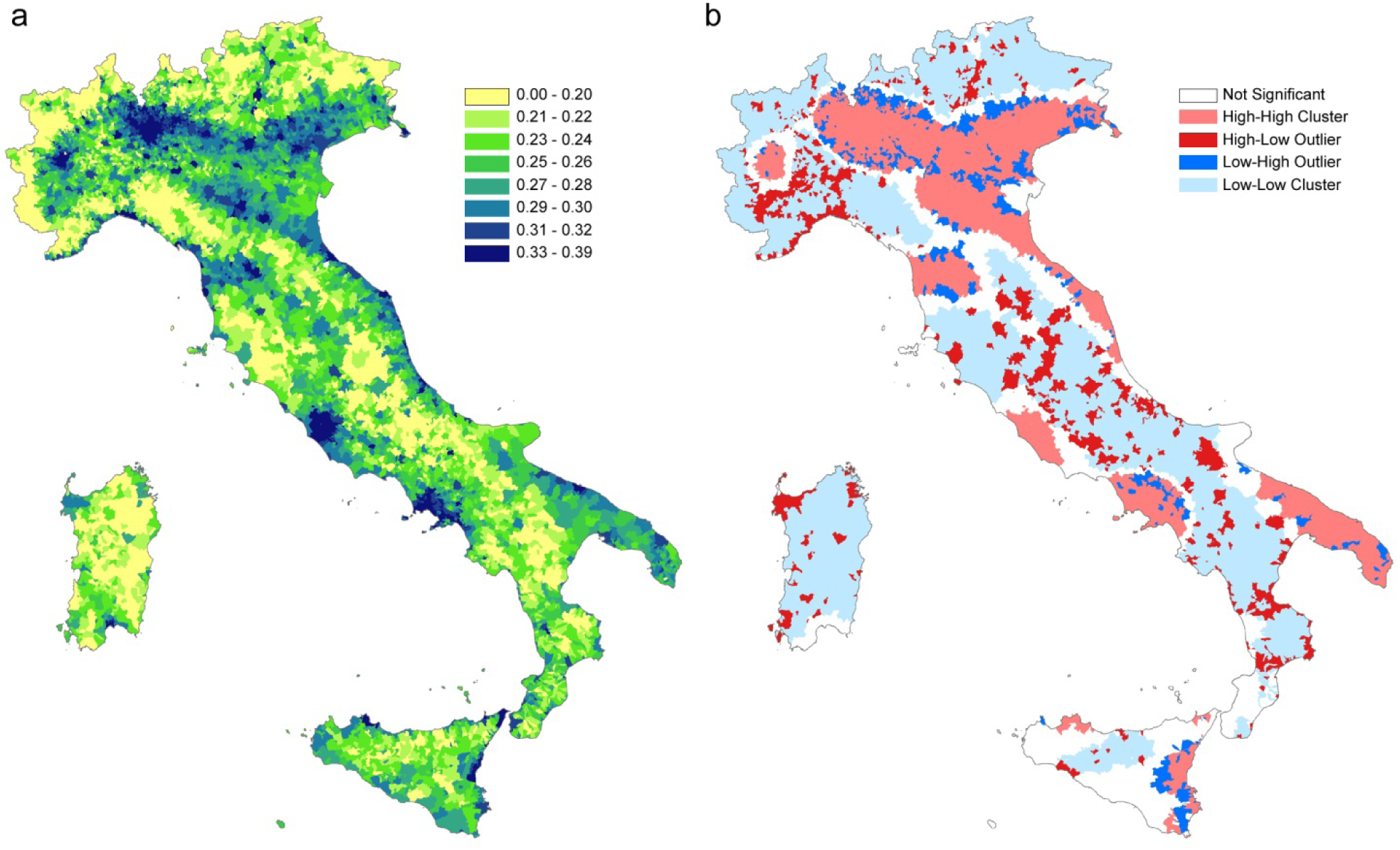
(a) β values per municipality (in legend quantile classification); (b) statistically significant hot spots in pink (municipalities with high βvalues in significant clusters), cold spots in light blue (clustered municipalities with low β values); in red municipalities with high β values and surrounded by municipalities with low β values; in blue municipalities with a low β value surrounded by municipalities with high values. Municipalities with not significant clustering or outliers are shown in white.

Three scenarios are evaluated as listed in Table 1: COVID-19 spreading at municipality level for the entire Italian territory between 26 February and 6 March (scenario 1); local spreading (during the first 21 days of the epidemic) in Lombardy, Abruzzi, and Basilicata regions (scenario 2); local spread in the Abruzzi region (during the first 14 days of the epidemic) considering each municipality as a possible origin of the infection (scenario 3).

#### Scenario 1

The number of infected provinces during the studied period increased from 29 to 92 and only 15 were still free from COVID-19 on 6 March. The number of infected people increased from 625 to 5699. Considering the results at national level, the model was able to model the number of cases as observed (Figure 6a), while the observed increase of infected provinces was more rapid than the estimated one (Figure 6b).

**Figure 6.**
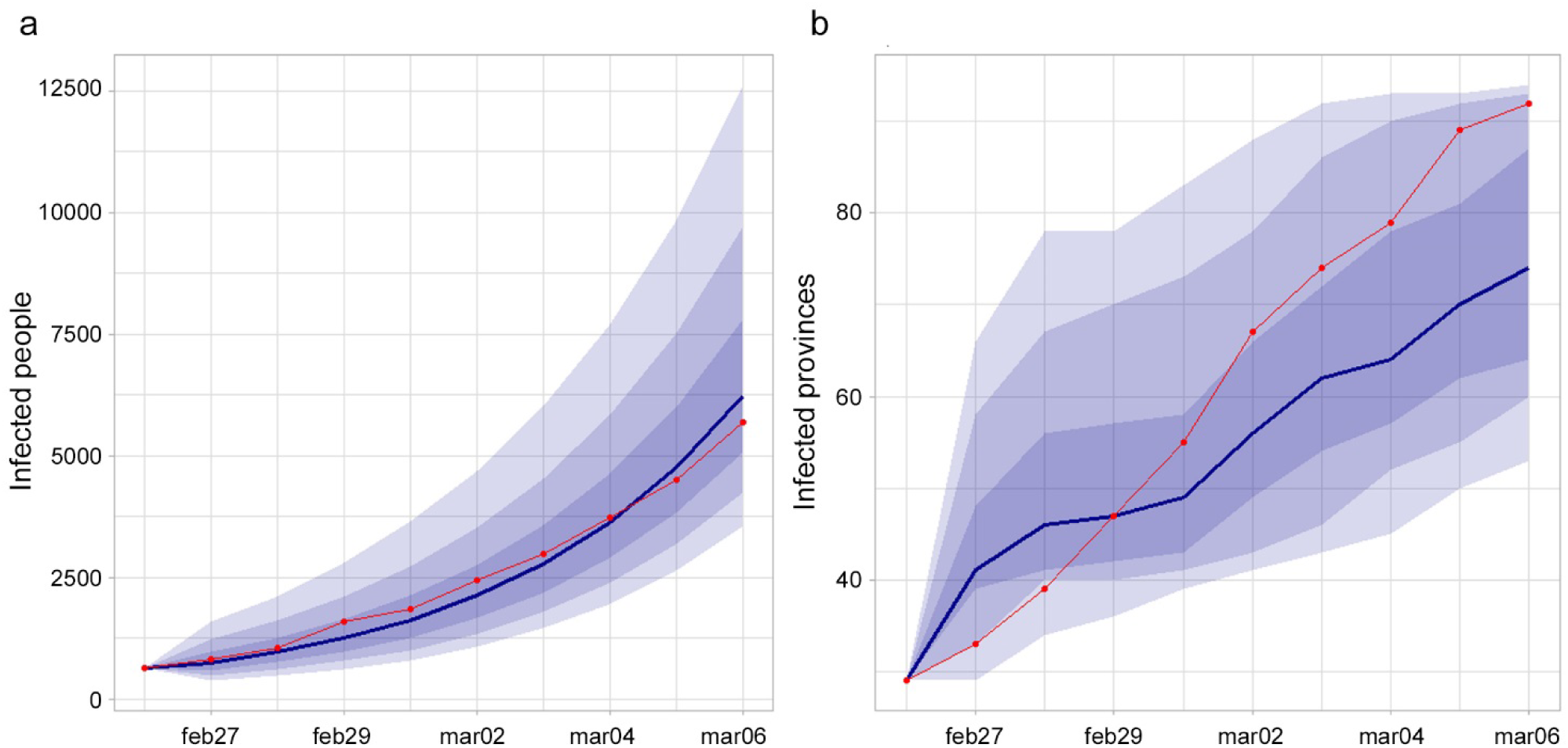
(a) Observed (red line) and estimated (blue line and 0.95, 080 and 0.50 CI) infected people in Italy. (b) Observed (red line) and estimated (blue line and 0.95, 080 and 0.50 CI) number of infected provinces.

The Pearson’s correlation coefficient between observed and estimated infected people (median value) at province level, at the end of the period was equal to 0.92.

Figure 7 shows the agreement between observed and estimated infected province on 6 March. Provinces’ color represents the estimated probability of being infected, red points indicates the observed infected province at the end of the period, green points represent the non-infected provinces at the end of the period, dashed provinces are the seeds at the beginning of the period.

**Figure 7.**
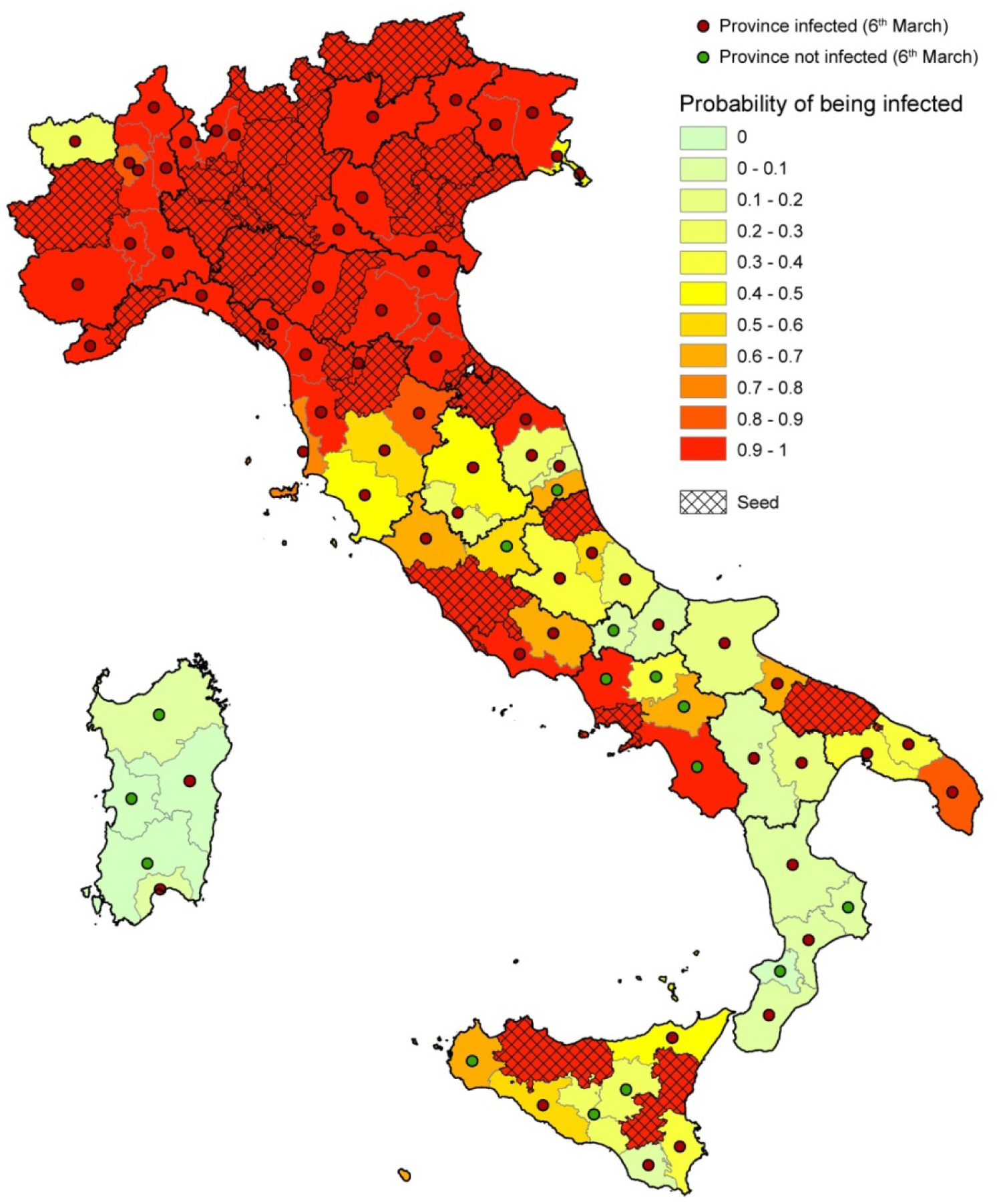
Results of the model. Provinces are colored based on the estimated probability of being infected; red points show the observed infected provinces at the end of the period (26 Feb-6 Mar); the green points represent the non-infected provinces at the end of the period; dashed provinces represent the seeds at the beginning of the period (26 Feb).

A threshold of p=5% on the percentage of simulations having at least one infected individual per province was chosen to evaluate the model performance in predicting the infection status of each province at the end of the period. The comparison with observed official status as of 6 March resulted in a number of True Positive (TP) = 85, True Negative = 6, False Positive (FP)= 9 and False Negative = 7. This leads to a Sensitivity = 92.4% and a Specificity = 40%, for a total Accuracy of 85%.

Figure 8 shows the comparison between observed (a) and estimated (b) infected people (upper 95% confidence interval when a province is turned into infected in the simulations) excluding the seeds.

**Figure 8.**
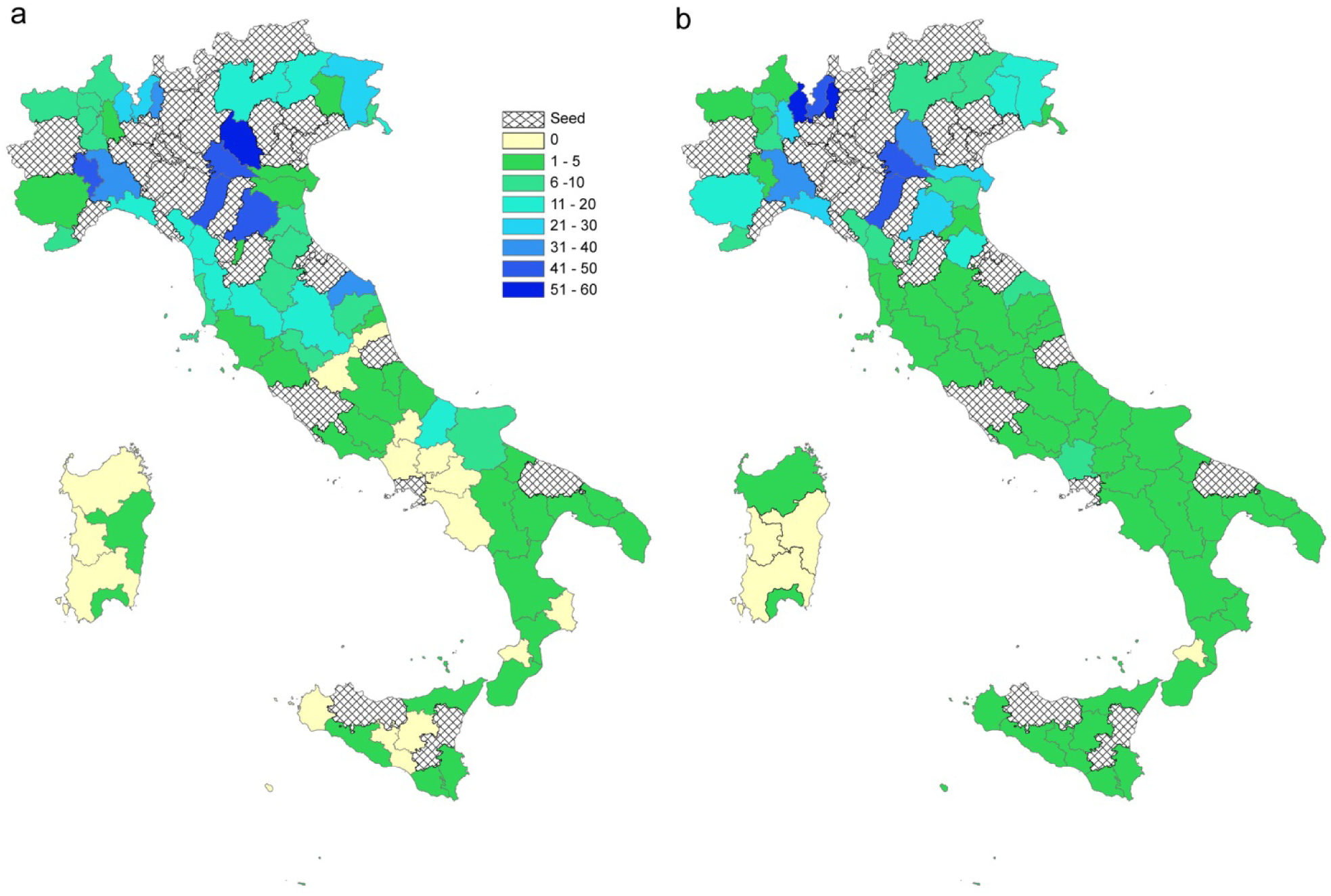
Results of the model. Comparison between observed (a) and estimated (b) infected people. The upper 95% confidence interval of the estimated infected people is used (when a province is turned into infected during the simulations the value of infected people is considered). Dashed provinces are the initial seeds.

As far as the seeding sites concern, Figure 9 shows the comparison between observed and estimated infected people in each seed province. For most of the cases the observed values fall into the 95% confidence interval of the estimates. Results are grouped in four panels depending on the magnitude of the observed COVID-19 epidemic.

**Figure 9.**
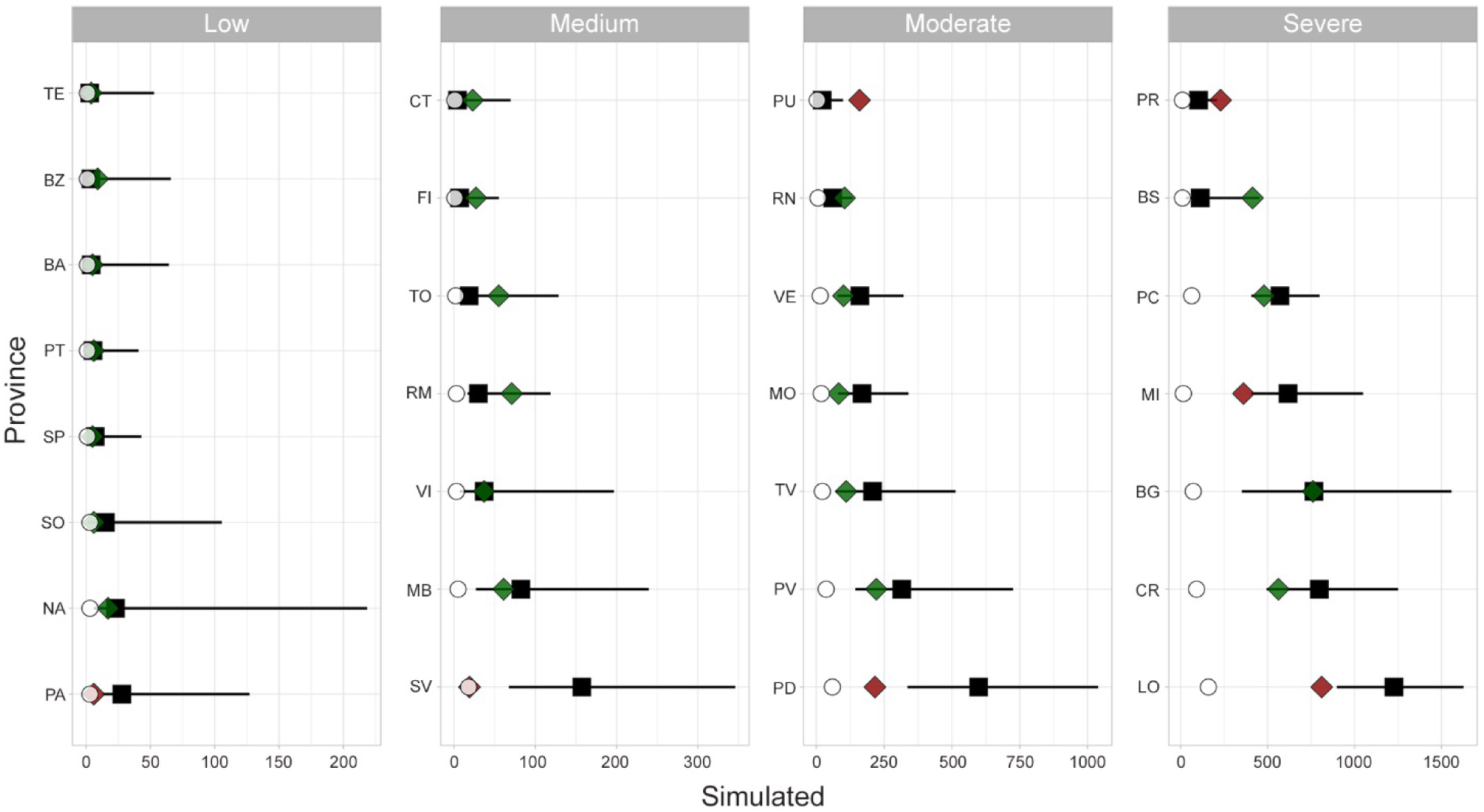
Infected people in seeding provinces on 6 March. Black squares represent median values, lines the 95%CI, green and red diamonds represent observed data within (green) and outside(red) the CI, whilst white dots represent the observed cases on 26 February. The panels order (from low to severe level of infected people) is used for displaying purpose.

#### Scenario 2

The differences observed among the spread patterns in high (Lombardy), medium (Abruzzi) and low (Basilicata) affected areas might be explained by different contact rate patterns and different commuting systems.

Figure 10 compares β values (a), degree measure (b) and estimated probability of being infected (c) for each municipality. Starting from one seed in each region (the first municipality notified as infected), for a time window of 21 days, the disease seems to follow different patterns: in Lombardy, where β values are more homogeneous, the disease extends in wideness; in Abruzzi region the disease spreads along the Adriatic coast, driven by the β parameter higher in this corridor; in Basilicata, the disease remains confined to the point of origin because the region has neither high β values or high connections.

**Figure 10.**
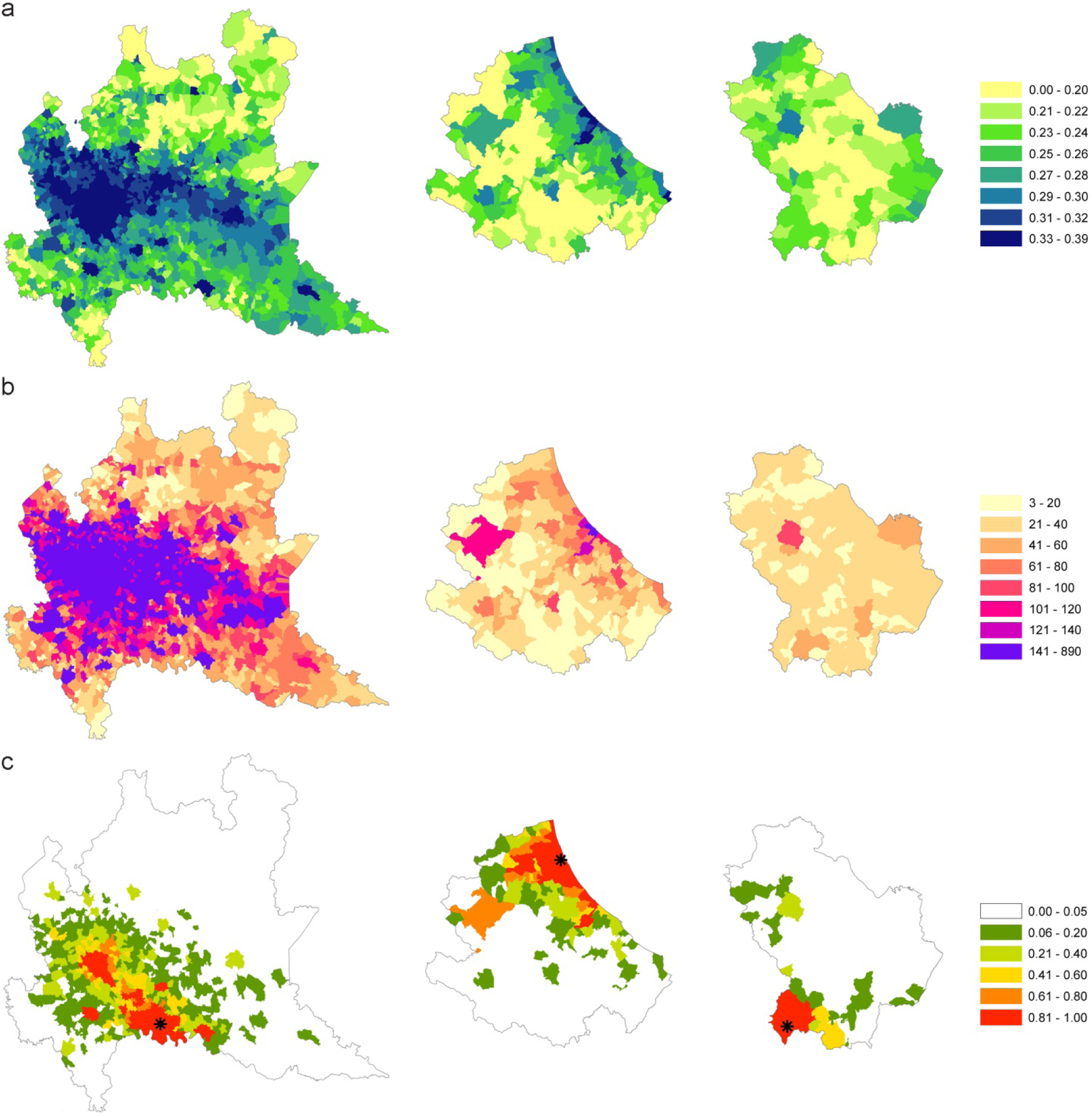
β values (a), degree measure (b) and estimated probability (c) of being infected for each municipality of high (Lombardy), medium (Abruzzi) and Low (Basilicata) affected areas.

Moreover, the differences among the three regions have been evaluated through the number of estimated infected people and infected municipalities (Figure 11a and b). The pattern of infected people is similar for Abruzzi and Lombardy (red and green line respectively), but different from Basilicata (green line). However, the differences are more evident between the three regions if we consider the number of infected municipalities that grows more rapidly in the case of Lombardy (Figure 11b). The speed with which the municipalities in Lombardy become infected is higher than in Basilicata and Abruzzi due to the connections underlying the commuting network (Figure 11c). It is noteworthy that Lombardy has 28% of municipalities with a degree greater than 140, while in Abruzzi and Basilicata regions 95% and 99% of municipalities respectively are below a degree of 60.

**Figure 11.**
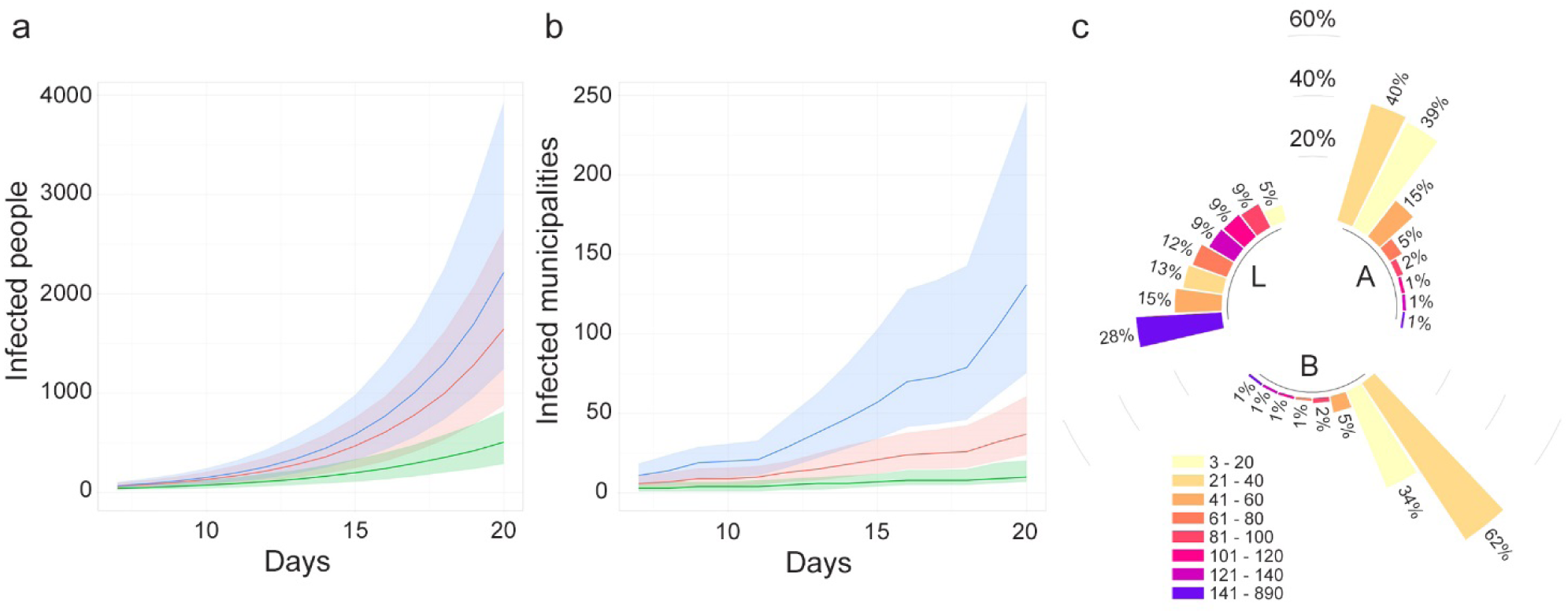
(a) Number of estimated infected people (median values and 95%CI) and (b) estimated infected municipalities (median values and 95% CI) when the epidemic starts in one seed and lasts 21 days. (c). Blue line represents Lombardy, red line is Abruzzi region and green line, Basilicata region. Degree distribution in the three region L=Lombardy, A=Abruzzi, B=Basilicata. Bars are in class percentage descending order.

#### Scenario 3

In this scenario, each municipality is considered in turn as seed. The vulnerability of Abruzzi region is calculated as the number of infected individuals (95th percentile) and infected municipalities that each municipality (seed) causes in the region (excluding itself) (Figure 12a and b). Figure 12c shows the ratio between the number of cases caused outside the municipality and the number of cases caused inside the municipality (x 100) that may be interpreted as the tendency of each municipality to act as a destination or origin of infection for the other territories.

**Figure 12.**
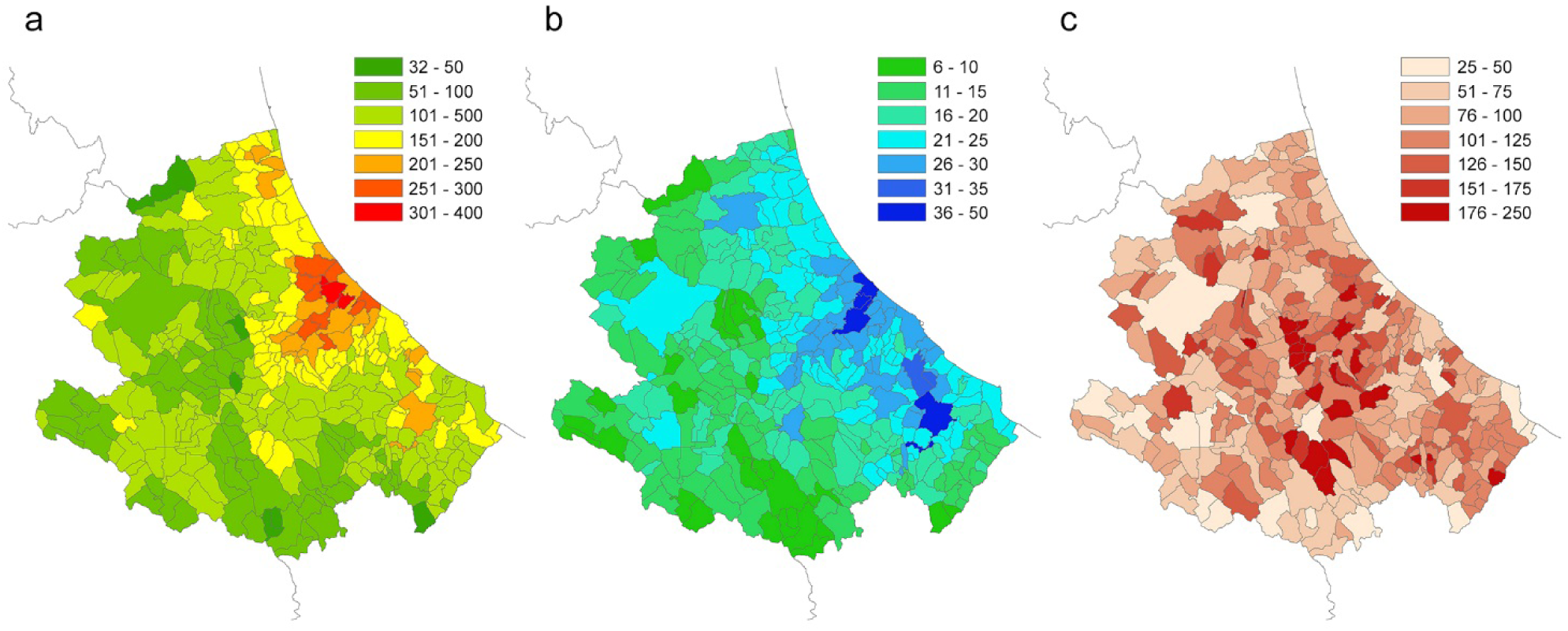
(a) Number of infected people (95th percentile) that each municipality causes in the region (excluding itself). (b) number of infected municipalities caused by each seed (95th percentile). (c) ratio between the number of cases caused outside the municipality (95th percentile) and the number of cases caused inside the municipality (95th percentile).

## Discussion and conclusions

Human mobility data have been largely used for modelling the spread of infectious diseases both at global [2,5,8,9,23,24] and national level [25–27]. A recent use of these data for modelling COVID-19 epidemic in Italy has been published by Gatto et al. [10] and Vollmer, et al. [6].

Data on commuting, defined as the daily local movements from home to work location or schools, have been used in the study of epidemiology of infectious disease at lesser extent.

In Italy the currently available data on commuting are census data, with the advantage of being structured, open source, and representative of the entire Italian population. Its update every 10 years does not seem to affect the spatial patterns of human mobility [10], thanks also to the stability of the production systems and persistence of attractive poles (schools, offices etc.) in the same places.

The use of data at a higher spatial resolution (municipality level) allows to highlight peculiar situations on which public health authorities may promptly intervene to control the spread of a disease. For this reason, we have introduced within a metapopulation model, driven by the commuting network, a municipality-based contact rate able to capture the variability between municipalities in terms of population density and commuting system.

We used a simple Susceptible-Infected model in which more importance was given to model the contact rate, neglecting the Exposed, Recovered and Dead compartments. This approach can be suitable in the first phase of an new epidemic (when population is fully susceptible). Although we have reduced the number of parameters of the epidemiological model from one side, we have introduced other variables to model the socio-demographic and commuting aspects, as detailed in Supplementary Information.

The revised calculation of the infection contact rate (β, based mainly on the resident population density, also incorporates the commuting component in each municipality, highlighting its characteristic of being an attractor of commuters or a displacer of workforces toward elsewhere. When areas with a high β values are contiguous and significantly clustered, disease permeability tends to be greater. Lombardy, Veneto and Emilia Romagna (situated in the northern Italy), which together made up 52% of all Italian cases (as of May 7, 2020), are effectively clustered with similar and high β values (Figure 5b).

The revised municipality-based β can be generalized to any other epidemic that responds to the assumptions made for its calculation.

The simulation model was applied considering three different scenarios. The first scenario (scenario 1), considering COVID-19 spreading at municipality level for the entire Italian territory between 26 February and 6 March, was used to assess the model accuracy.

At national level, the model estimates the trend of infected people similarly to the observed trend of cases (Figure 6a), despite the introduction of the β variability. As far as the number of provinces involved concerns, the estimates are more variable.

In particular, the model estimates a growth rate of the infected provinces lower than the observed one (Figure 6b). This might be due to the uncertainty about the real number of provinces already infected at the beginning of the period. If a higher number of infected provinces at the beginning had been considered, the outcomes of the model would have been more similar to the observed ones.

Considering the capacity of the model to correctly classify a province as infected at the end of the observation period, despite a global accuracy of 85%, the model failed in classifying 16 provinces. However, the misclassification is due not only to the model capacity, but also to the influence of uncontrolled factors such as errors in the observed data, timing in notification of cases, ability to identify the disease, containment local measures, long distance displacement of people from infected areas to non-infected areas.

As for the false negative (FN) provinces, the model failed in identifying as infected, provinces in which the number of observed cases at the end of the study period was actually very low (from 1 to 3); only one province out of 7 never turned out infected in any simulation. Furthermore, most of the FN provinces are located in southern Italy, an area which was affected by the massive return of university students from the North, after all schools were closed. The 9 false positive (FP) provinces, were officially detected as positive a few days after the considered period.

The outcomes of the model were compared with the observed number of COVID-19 cases for the initially infected provinces, grouped by different virus circulation level (from low to high). For some Provinces, such as Palermo (PA), Savona (SV), Padua (PD), Milan (MI) and Lodi (LO) the model overestimates the cases, whereas in few others, like Pesaro-Urbino (PU) and Parma (PR) the number of cases was underestimated (Figure 9). One of the possible explanations for this disagreement may be found in the application of control measures by local authorities, which anticipated the measures of the central government. On the other hand, the model was able to estimate quite precisely the number of cases in several provinces (either at low or high virus circulation level). Among these, Bergamo (BG) province was one of the Italian provinces that suffered more for the COVID-19, with more than 10,000 cases and almost 3,000 deaths. This would lead to think that in that province the control measures were not strictly applied at the very beginning of the epidemic. It is important to note that the limitation of the model in correctly estimating the magnitude or the extent of the epidemic also depends on the difficulty of including in the model the establishment of community (hospitals, health care settings, working) or household clusters of infection.

The second scenario (scenario 2) was developed to explore and compare the spread pattern in three different regions. The chosen regions (Lombardy, Abruzzi and Basilicata) are characterised by different epidemiological conditions. When the outcomes for the three regions are compared, the population density and level of industrialization must be taken into account. The mobility of workforces in Lombardy, which is one of the major economic driving areas in Europe, is much greater than in the other two regions. The differences among these regions are more evident when the increase of the number of infected municipalities is considered (Figure 11b), being the infection spreading across municipalities directly linked to the connections underlying the commuting network. Indeed, in case of Lombardy about the 50% of the municipalities are connected to more than 100 municipalities (against a 3% in Abruzzi and Basilicata regions – Figure 11c).

In the last scenario (scenario 3) the local spread in the Abruzzi region is estimated (during the first 14 days of the epidemic) considering each municipality as a seed for simulation. This scenario has the purpose of identifying those municipalities more vulnerable to the virus introduction and those playing a major role in spreading the infection. The vulnerability of Abruzzi region is calculated as the number of individuals that each municipality (seed) causes in the region and the number of infected municipalities (Figure 12a and b). In addition, the ratio between the number of cases caused outside the municipality and the number of cases caused inside the municipality may provide a useful hint about the risk category (capability to infect rather than become infected, shown in Figure 12c) of each municipality. The obtained maps, at municipality level, provide the decision makers with useful information on where mobility restriction measures should be focussed to have the strongest effects on transmission reduction. The highest vulnerability values can be observed in areas with commercial hubs, close to the highest populated city of the region, Pescara (Figure 12b, the darker blue area on mid-coastal line) and the most industrial area of the region, in the Sangro Valley (Figure 12b, in the southern of Abruzzi region), where many medium and big factories are present.

Our approach, therefore, provides decision-makers with useful geographically detailed metrics to evaluate those areas at major risk for infection spreading and for which restrictions of human mobility would give the greatest benefits, especially in the first phase of the epidemic. It can provide risk maps on which health administration can modulate the application of strong lockdown measures, evaluating in advance the effects on reducing the spread of the infection.

This approach is particularly useful not only in the beginning of the epidemic but also in the last phase, when the risks deriving from the gradual lockdown exit strategies must be carefully evaluated. In fact, the major risk in this latter phase is the re-insurgence of infection transmission through a progressive re-opening of the productive systems. The analysis of daily human mobility patterns for working reasons is clearly providing a well detailed picture of the areas and productive systems more at risk of sustaining a restart of the epidemic.

## Data Availability

all data in the paper are open data

## Notes

### Competing Interest Statement

The authors have declared no competing interest.

### Funding Statement

no external funding was received

